# DNA damage, nucleolar stress and dysregulated energy metabolism as mechanisms of multimorbidity in cardiovascular disease

**DOI:** 10.1101/2023.02.22.23286318

**Authors:** Kristina Tomkova, Marius Roman, Adewale S Adebayo, Sophia Sheikh, Syabira Yusoff, Melanie Gulston, Lathishia Joel-David, Florence Y Lai, Antonio Murgia, Bryony Eagle-Hemming, Hardeep Aujla, Tom Chad, Gavin D Richardson, Julian L Griffin, Gavin J Murphy, Marcin J Woźniak

## Abstract

**Background:** People with two or more underlying chronic conditions (multimorbidity) are more susceptible to adverse events following surgery. This study used -omics methods to compare samples from people with or without multimorbidity in a cardiac surgery cohort.

**Methods and Results:** Measurements of the myocardial transcriptome and metabolites involved in energy production were performed in 53 and 57 sequential participants, respectively. Untargeted analysis of the metabolome in blood and myocardium was performed in 30 sequential participants. Mitochondrial respiration in circulating monocytes was measured in 63 participants. Ninety-eight of 144 participants (68%) had multimorbidity. Three major processes were affected by multimorbidity: innate immune response, DNA damage and associated epigenetic changes, and mitochondrial energy production. The innate immune response was upregulated in multimorbidity and most of the included comorbidities. The DNA damage, epigenetic changes, and aspects of mitochondrial function were specific for multimorbidity. Histone 2B, its ubiquitination enzymes and AKT3 were upregulated in the multimorbid group, supporting our hypothesis of senescence-like changes in multimorbidity. In addition, senescence-associated secretory phenotype analytes, IL-1β, its receptor, GM-CSF and fractalkine increased with the number of accumulating comorbidities. DNA damage was confirmed by independent immunohistochemistry experiments, which also identified nucleolar instability as more prominent in the multimorbid myocardium.

**Conclusions:** Multimorbidity in people with cardiovascular disease is characterized by biological aging processes that are known to increase susceptibility to metabolic stress. These present novel therapeutic targets for organ protection interventions.

## Introduction

Multimorbidity, defined as the presence of two or more comorbid conditions, affects over 50 million people in the European Union. (Navickas et al., 2016) Multimorbidity is associated with frailty, increased susceptibility to stressors, functional decline after acute illness, and increased use of healthcare resources. (Fortin et al., 2007) In the UK, multimorbidity is projected to affect two-thirds of adults aged over 65 years by 2035. (Kingston et al., 2018) As research traditionally focuses on diseases in isolation, the mechanisms underlying multimorbidity are poorly understood. (Fortin et al., 2007) Moreover, people with multimorbidity are often excluded from clinical trials of interventions and are therefore managed in the absence of high-quality evidence. Improving the lives of all people with multimorbidity is considered a national and global health research priority. (2018)

Two-thirds of people with cardiovascular disease have multimorbidity. They suffer from larger infarct sizes after acute coronary syndrome (ACS), worse prognosis in heart failure, and attenuated effectiveness of organ protection strategies. (Rucker & Joseph, 2022; Ruiz-Meana et al., 2020) In cardiac surgery, multimorbidity affects 86% of people >65 years, (Forman et al., 2018) and is associated with a threefold increase in mortality. (Stirland et al., 2020) As the population ages, the health burden attributable to cardiovascular disease in the setting of multimorbidity will increase. Recent reviews suggest chronic inflammation as the major factor driving the increased susceptibility to metabolic stress in multimorbidity. (Skou et al., 2022) It is likely responsible for epigenetic changes, the disruption of energy production, oxidative stress and senescence. The research into each of these processes usually involves isolated cells and animal models. However, no cell-based or animal model can replicate multifactorial multimorbidity in humans. This study fills this gap. We decided on a hypothesis-free approach using unbiased approaches including untargeted transcriptomics and metabolomics to identify specific processes in people with cardiovascular disease and multimorbidity. The identified processes were further verified in targeted assays including direct measurement of mitochondrial respiration in circulating monocytes.

## Methods

### Study design

A prospective observational case-control study to identify the role of epigenetic regulation of genes responsible for energy metabolism and mitochondrial function in the obesity paradox in cardiac surgery (ObCARD) was approved by The East Midlands – Nottingham 1 Research Ethics Committee. All participants provided written informed consent. The study protocol was registered at clinicaltrials.gov; NCT02908009. A primary analysis of this data has been reported previously. (Adebayo et al., 2022) This report is a secondary analysis, pre-specified in the study protocol. The study is reported as per the STrengthening the Reporting of Observational studies in Epidemiology (STROBE) statement. The STROBE checklist is included in the supplementary material (**Table S1**). The study adhered to the principles outlined in the Declaration of Helsinki.

### Study cohort

Adults (>16 years) undergoing coronary artery bypass grafting with or without valve surgery. Exclusions included pre-existing paroxysmal, persistent or chronic atrial fibrillation, pre-existing states likely to have hyperinflammatory phenotypes (sepsis undergoing treatment, acute kidney injury within five days, autoimmune diseases, chronic infection, congestive heart failure), ejection fraction <30 %, pregnancy and in a critical pre-operative state (stage 3 AKI (Kidney Disease: Improving Global Outcomes (KDIGO) Acute Kidney Injury Work Group, 2012) or requiring inotropes, ventilation or an intra-aortic balloon pump). Emergency or salvage procedures were also excluded. The study was designed to be hypothesis-generating, and no sample size was specified.

Multimorbidity was defined as per the Academy of Medical Sciences definition of two or more long-term (> 1 year duration) conditions. (Quality standard [QS153], 2017) In addition, in sensitivity analyses we also used the *complex multimorbidity* definition where two or more *physiological systems* were affected. (I. S.-S. Ho et al., 2021) Obesity was defined as BMI > 32, based on our previous analysis. (Adebayo et al., 2022) Angina/MI was defined as Canadian Cardiovascular Society (CCS) angina grade II or higher or previous myocardial infarction. Recruitment was determined by the simultaneous availability of a consented patient undergoing surgery, clinical research staff, and laboratory research staff available to undertake analyses of fresh tissue and cells. Patients were recruited consecutively based on this availability.

### Sampling

Right atrial biopsies were collected in a standardized manner from the right auriculum before cannulation for cardiopulmonary bypass and were immediately snap-frozen in liquid nitrogen. Blood samples were processed within two hours of collection for the respiration analysis. The remaining plasma samples were frozen and stored at −80C.

### Measures taken to reduce bias

Selection bias was mitigated by the recruitment of sequential patients. Detection bias was mitigated by blinding laboratory staff who analyzed atrial biopsies and blood samples.

### RNA isolation and sequencing

RNA was isolated from 20mg of tissue using Isolate II RNA mini kit (Bioline, London, UK). Sample quality was assessed using the RNA Screentape assay on the Agilent TapeStation 4200. Only samples with RNA integrity numbers equal to or greater than eight were sequenced.

Library preparation and sequencing were carried out in two batches by Source BioScience (Nottingham, UK). The stranded total RNA libraries were prepared in accordance with the Illumina TruSeq stranded total RNA sample preparation guide with Ribo-Zero human/mouse/rat for Illumina paired-end multiplexed sequencing. The libraries were validated on the Agilent Bioanalyzer 2100 to check the size distribution of the libraries and on the Qubit high sensitivity to check the concentration of the libraries. Sequencing was performed using 75bp paired-end chemistry on HiSeq 4000 with the TruSeq stranded total RNA human kit.

### Metabolomics

Untargeted metabolomics was performed by Metabolon, Inc. (USA). All samples passed appropriate quality controls.

For targeted metabolomics, a panel of 144 cellular metabolites involved in mitochondrial function and energy metabolism were analyzed on a Thermo Qunativa interfaced with a Vanquish LC as previously described. (Charidemou et al., 2019) In brief, tissue was extracted using a modified Folch extraction into chloroform/methanol (2:1 600 ul per 50 mg of tissue, followed by 200 ul of water, 200 ul of chloroform, repeated once). For nucleotides and acyl-CoA derivatives, one-half of the aqueous extract was dissolved in 150 µl of 70:30 acetonitrile:water containing 20 µM deoxy-glucose 6 phosphate and 20 µM [U–13C, 15N] glutamate. The resulting solution was vortexed, sonicated and centrifuged. Chromatography consisted of a strong mobile phase (A) was 100 mM ammonium acetate, and the weak mobile phase was acetonitrile (B) and the LC column used was the ZIC-HILIC column from Sequant (100 mm × 2.1 mm, 5 µm).

For amino acids and TCA cycle intermediates, aqueous extracts were reconstituted in 50 μl of 10 mmol/l ammonium acetate in water before TCA cycle intermediates were separated using reversed-phase liquid chromatography on a C18-PFP column (150 mm × 2.1 mm, 2.0 μm; ACE). For chromatography on the UHPLC system, mobile phase A was 0.1% formic acid in water, and mobile phase B was 0.1% formic acid in acetonitrile. Mass transitions of each species were as follows (precursor > product): D5-L-proline 121.2 > 74.2; D8-L-valine 126.1 > 80.2; D10-L-leucine 142.0 > 96.2; L-glutamate [M] 148.0 > 84.2; L-glutamate [M+1] 149.0 > 85.2; L-glutamate [M+6] 154.1 > 89.1; citrate 191.0 > 111.0; citrate [M+1] 192.0 > 112.0; citrate [M+2] 193.0 > 113.0; citrate [M+3] 194.0 > 114.0; citrate [M+4] 195.0 > 114.0; citrate [M+5] 196.0 > 115.0; citrate [M+6] 197.0 > 116.0. Collision energies and radio frequency (RF) lens voltages were generated for each species using the TSQ Quantiva optimization function.

### Mitochondrial respiration measurements

were performed in circulating monocytes isolated from pre-operative blood samples using the Histopaque method. Oxygen consumption (OCR) and extracellular acidification rates (ECAR) were measured in the absence or presence of 4µM oligomycin, 2µM FCCP or 2µM rotenone and 2µM antimycin A (Merck, UK) with Seahorse XFe24 analyzer (Agilent Technologies, USA) in 200,000 cells/well. To assess the effect of multimorbidity on glycolysis, OCR was measured in the presence of glucose or pyruvate as substrates. The respiratory control ratio was calculated as described in Hill et al.: RCR_max_=(FCCP-Antimycin)/(Oligomycin-Antimycin); RCR_basal_=(Basal-Antimycin)/(Basal-Antimycin) (Hill et al., 2012) For each sample at least three parallel measurements were performed. Samples with poor background respiration or insufficient measurements were removed from the analysis.

### Cytokine and chemokine panel

was assessed by measuring a panel of 71 cytokines/chemokines in plasma samples collected before and 24 hr after surgery by Eve Technologies (Canada). All included samples passed quality control.

### Immunohistochemistry

was performed on cardiac biopsies. Tissue samples were frozen in OCT (Cell Path, UK) and sliced using Leica CM1520 cryostat (Leica Microsystems, UK) at 10-micron thickness. The slices were fixed in 10% neutral buffered formalin (Merck, UK) and permeabilized in ethanol (50%, 70%, and 100% EtOH). Unspecific binding was blocked with 1% BSA (Merck, UK). Samples were labeled with primary antibodies against nucleolin (rabbit polyclonal, abcam, UK), fibrillarin (rabbit polyclonal, abcam, UK), γH2AX (rabbit polyclonal, abcam, UK) or cardiac troponin T (mouse clone 1C11, abcam, UK). All primary antibodies were used at 1:300 dilution in Co-Detection Antibody Diluent (Advanced Cell Diagnostics, USA). The primary antibodies were detected with secondary Alexa Fluor 568 goat-anti-rabbit (Invitrogen, UK) and Alexa Fluor 488 goat-anti-mouse (Invitrogen, UK) antibodies at 1:200 dilution. Nuclei were labelled with DAPI (Thermo Fisher Scientific, UK). The sections were treated with Prolong Gold Anti-fade media (Invitrogen, UK), and visualized using an inverted Zeiss Axio Observer Z1 microscope equipped with Colibri 2 LED illumination, Plan-Apochromat 63x/1.40 oil objective and ORCA-Flash4.0 CMOS camera (Hamamatsu Photonics, Japan). For each patient’s sample, three slices were prepared, and approximately 20 images were collected per slice (400 – 500 cells). Images were analyzed using FIJI ImageJ distribution. Only the nuclear fraction of nucleolin staining and nuclear fibrillarin-positive particles were considered in the analysis.

### Quantitative real-time PCR

was used to estimate ribosomal DNA (rDNA) copy numbers. The genomic DNA was extracted from cardiac biopsies using a commercial kit following the manufacturer’s instructions (Genomic-tip 20/G and Genomic DNA Buffer Set, QIAGEN, UK). Specific primers for amplification of 5S, 5.8S, 18S, and 28S rDNAs were designed using NIH Primer-BLAST tool (Ye et al., 2012) and synthesized by Merck (UK). As an endogenous PCR control, primers for the β-2-Microglobulin (B2M) gene were used. Primer sequences: rDNA 5S forward TCGTCTGATCTCGGAAGCTAA, reverse AAGCCTACAGCACCCGGTAT; rDNA 5.8S forward GAGGCAACCCCCTCTCCTCTT, reverse: GAGCCGAGTGATCCACCGCTA; rDNA 18S forward: AGCCTGAGAAACGGCTACCA, reverse: GGTCGGGAGTGGGTAATTTGC; rDNA 28S forward: CTCCGAGACGCGACCTCAGAT, reverse: CGGGTCTTCCGTACGCCACAT; B2M forward primer TGCTGTCTCCATGTTTGATGTATCT, reverse primer TCTCTGCTCCCCACCTCTAAGT. For each 20μl PCR reaction, 6ng of genomic DNA was combined with 10μl of PowerUp SYBR Green Master Mix (Thermo Fisher Scientific, UK), 1μl of specific primers at 10μM, and nuclease-free water. All reactions were set up in triplicates and measured using a Rotor-Gene Q qPCR machine (Qiagen, UK). Melt curves were produced for all measurements to confirm the presence of a single PCR product. The data were analyzed using the Rotor-Gene Q 2.1.0.9 software (Qiagen, UK).

### Data processing and statistical analysis

Unless indicated otherwise, data analysis was performed with R Statistical Computing software version 4.2.2, and plots were prepared with the ggplot2 R package. (R Core Team, 2022; Wickham, 2009)

*Transcriptomics* Sequencing data were quality-checked with FastQC v0.11.5, quantified with Salmon v1.21 (Patro et al., 2017) after indexing with a decoy and annotated with Ensembl v100. Gene quantities were normalized to length-scaled transcripts per million and filtered for low quantities before downstream analysis using the limma-voom model (Ritchie et al., 2015) with empirical Bayes moderation. Sample groups were analyzed with the sequencing batch added to the model as a variable. The false discovery rate was set at 5%. Interactions within networks were visualized with Cytoscape. (Shannon et al., 2003)

Weighted gene correlation network analysis (Langfelder & Horvath, 2012) was carried out to cluster differential genes into smaller modules with the soft r-squared set at >0.8 and grey modules excluded. Each module formed was subsetted from the adjacency matrix and exported as edge files for visualization in Cytoscape.

To directly analyze the well-known signaling pathways in the data, a gene-set analysis was carried out on the filtered transcriptome data using camera (Wu & Smyth, 2012) with Reactome and Gene Ontology annotations for transcripts and non-coding RNA, respectively, with false discovery rate set at 5%.

### Metabolomics

Untargeted metabolomic profiles were processed by Metabolome Inc. For targeted metabolite analysis, the peak area ratio of metabolites was obtained by integration within vendor software (Xcalibur QuanBrowser, Thermo Scientific, Hemel Hempstead, UK) and compared with isotopically labeled standards for quantification. Data were pre-processed by removing constant-value features, replacing zeros and missing values with half the smallest value in the entire dataset and removing extremely low relative standard deviation using Metaboanalyst v5.0. (Xia et al., 2009) The processed data was log-transformed and mean-normalized. Pairwise comparisons of sample groups were carried out using a t-test. Metabolite set enrichment analysis was performed with Metaboanalyst v5.0.

### Multiomics analyses

of RNA and metabolite were combined using sparse Partial Least Squares (sPLS) models using mixOmics version 6.22.0. (Lê Cao et al., 2009) Canonical correlation patterns and association networks derived from the components were then used to infer the relationship between genes and metabolites. The network analysis was visualized with Cytoscape.

## Results

### Study cohort

Out of 1,021 people screened, 151 were recruited for this study. The reasons for exclusion are detailed in **Figure 1A**. Six participants withdrew after the consent, and one was excluded for protocol deviation. Samples from 144 participants were analyzed. Analysis of the metabolome was performed in samples from 30 sequential participants. Measurements of transcriptome and metabolites involved in energy production were performed in 53 and 57 sequential participants, respectively, as previously described. (Adebayo et al., 2022) Mitochondrial function in circulating monocytes was performed in 70 participants. Targeted validation was performed in samples from 29 – 48 participants and included analysis of senescence-associated secretory phenotype proteins in plasma, as well as estimations of ribosomal DNA copy numbers, DNA damage and nucleolar structure in the myocardium (**Figure 1A, B**).

**Figure 1.**
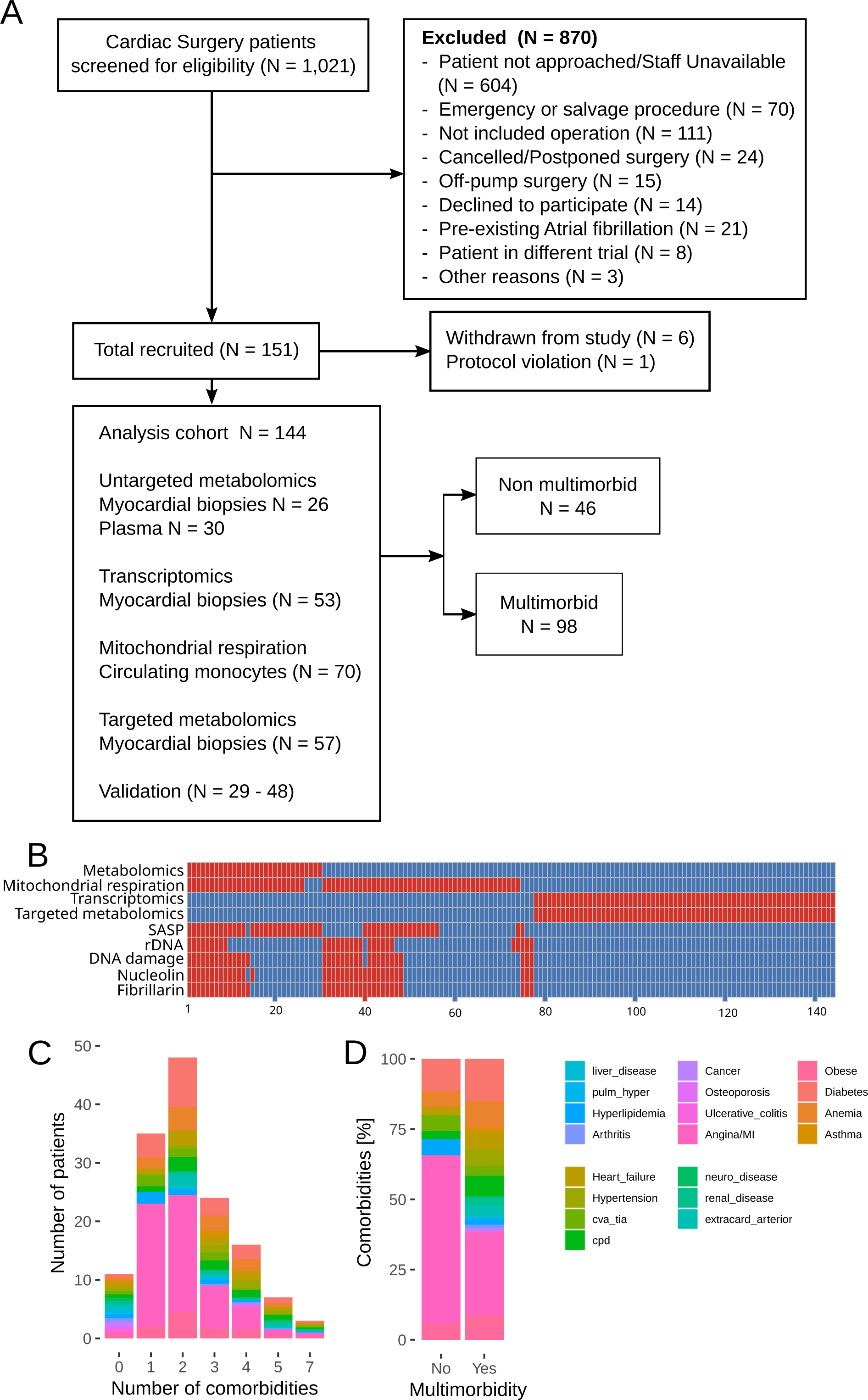
Cohort characteristics. **A** – CONSORT diagram; **B** – Breakdown of samples between the analyses. **C** – Comorbidities distribution in participants without or with a specific number of comorbidities; **D** – Comorbidities distribution in participants without and with multimorbidity.

Multimorbidity was defined as the presence of two or more of nineteen chronic conditions (**Table 1**). Ninety-eight participants (68%) suffered from at least two comorbidities (**Figure 1A**). The number of comorbidities per patient ranged between none (11 participants) and seven (three participants) and peaked at two comorbidities per patient (N=48, **Figure 1C, D**). Hypertension (12% vs 0%), angina or previous MI (49% vs 9%), heart failure (12% vs 1%), extracardiac arteriopathy (8% vs 0%), diabetes (30% vs 3%), obesity (24% vs 3%), chronic obstructive pulmonary disease (COPD; 15% vs 1%) and anemia (19% vs 1%) were enriched in the multimorbid group (**Table 1**). Post-surgery, the multimorbid group experienced higher lactate levels, lower mean PiO2/FiO2 ratio indicative of lung injury **(Figure S1)**, and 11 out of 12 cases of Acute Kidney Injury (**Table S2**).

**Table 1.**
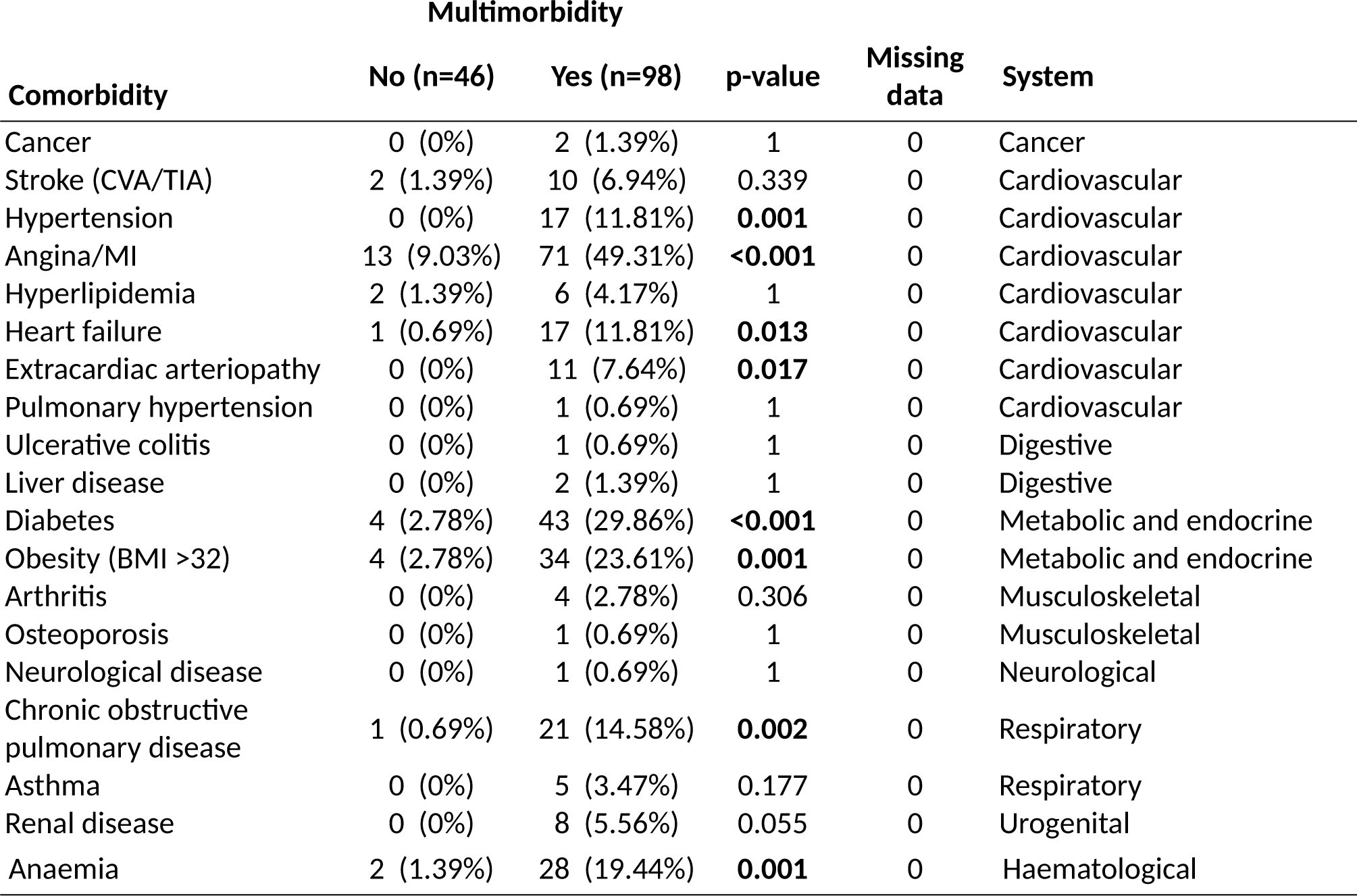
Distribution of comorbidities between the groups.

### Metabolite analyses

Untargeted metabolomics was performed in myocardial biopsies and pre-surgery plasma samples. Myocardial biopsies from four participants were of insufficient quantity (<30 mg) and were not included in the analysis.

The myocardial biopsy data set comprises 919 compounds, 820 of known identity and 99 of unknown structural identity. The plasma data set comprises 1316 compounds, 1046 of known identity and 270 of unknown structural identity (**Figure S2A-B**). All samples were comparable in numbers of known, unknown and undetected metabolites (**Figure S2A**). The most numerous metabolites were lipids (376 in biopsies and 483 in plasma), followed by amino acids (164 in biopsies and 216 in plasma, **Figure S2B**). There was no difference in the number of detected myocardial metabolites and their average peak area between participants with multimorbidity and without. The number of metabolites detected in plasma was significantly higher in multimorbid participants, although their average peak area was not different (**Figure S2C**).

There was a clear separation between multimorbid and non-multimorbid plasma samples in the principal component analysis. However, such distinction was not apparent for myocardial samples (**Figure 2A**). Differential expression analysis identified 29 upregulated and eight downregulated metabolites in myocardial biopsies. One hundred and nine metabolites were upregulated and 15 downregulated in plasma (**Figure S2D**). Since none of these metabolites passed the false discovery rate adjustment (**Tables S3** and **S4**), we performed KEGG metabolite set enrichment analysis using the whole dataset. The analysis of myocardial biopsies did not return any significant pathways. Plasma metabolites significantly (FDR<0.01) enriched Caffeine metabolism, Primary bile acid biosynthesis, Arginine and ornithine metabolism, Aminoacyl-tRNA biosynthesis, Glutamine and glutamate metabolism and Selenocompound metabolism (**Figure 2B**). Caffeine metabolism, Aminoacyl-tRNA biosynthesis and Primary bile acid biosynthesis were enriched with 9, 6 and 4 metabolites that differed (*p* < 0.05) between multimorbid and non-multimorbid groups. Levels of caffeine and its breakdown products, as well as bile acid intermediates, were higher in the multimorbid group, which indicates decreased expression or activity of cytochrome p450 enzymes, methylxanthine N1-demethylases, or choloylglycine hydrolase (**Figure S3A, B**). Levels of amino acids that enriched the other pathways were lower in the multimorbid group (**Figure 2B** and **Table S4**).

**Figure 2.**
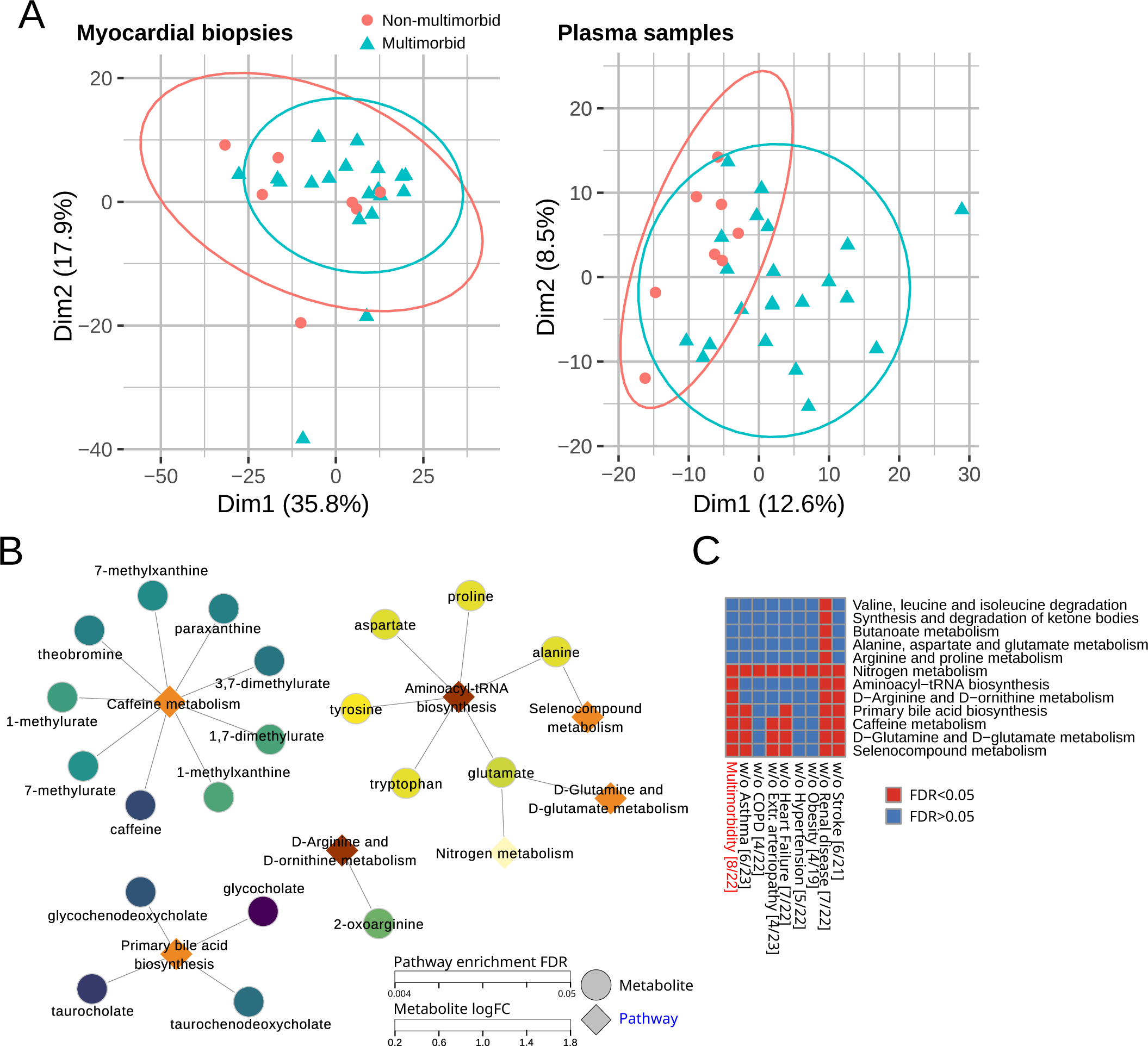
Metabolite analysis. **A** – Principal component (1 and 2) plots for the detected metabolites in the myocardial biopsies and plasma. **B** – KEGG metabolite set enrichment analysis in plasma samples with enriched metabolites. The color scales indicate metabolite log fold change (with positive number indicating upregulation in multimorbidity) or pathway enrichment p-value. **C** – KEGG metabolite set enrichment analysis in plasma in complex multimorbidity and in reduced datasets to test the influence of each comorbidity and anti-diabetic medications. Red squares indicate significantly enriched pathways for specific dataset. Numbers in square brackets indicate the number of non-multimorbid/multimorbid patients.

To assess the influence of each comorbidity on multimorbidity in our dataset, we performed KEGG metabolite set enrichment analysis without patients with that comorbidity. **Figure 2C** shows that Caffeine metabolism was affected by COPD, hypertension and Obesity; Primary bile acid biosynthesis was also affected by extracardiac arteriopathy; and Aminoacyl-tRNA biosynthesis was in addition affected by asthma. We were not able to analyze multimorbidity without patients who had diabetes, anemia, or who were on diabetic medications. Groups of patients with or without complex multimorbidity were identical to the primary multimorbidity classification.

### Transcripts analysis

The transcriptomics analysis was performed on myocardial biopsies, and a summary of the data set has been published previously. (Adebayo et al., 2022) Multimorbidity did not have a major effect on global gene expression patterns, as indicated by the principal component analysis (**Figure 3A**). The differential expression analysis detected 854 transcripts (**Table S5**). Most upregulated transcripts encoded immunoglobulin chains.

**Figure 3.**
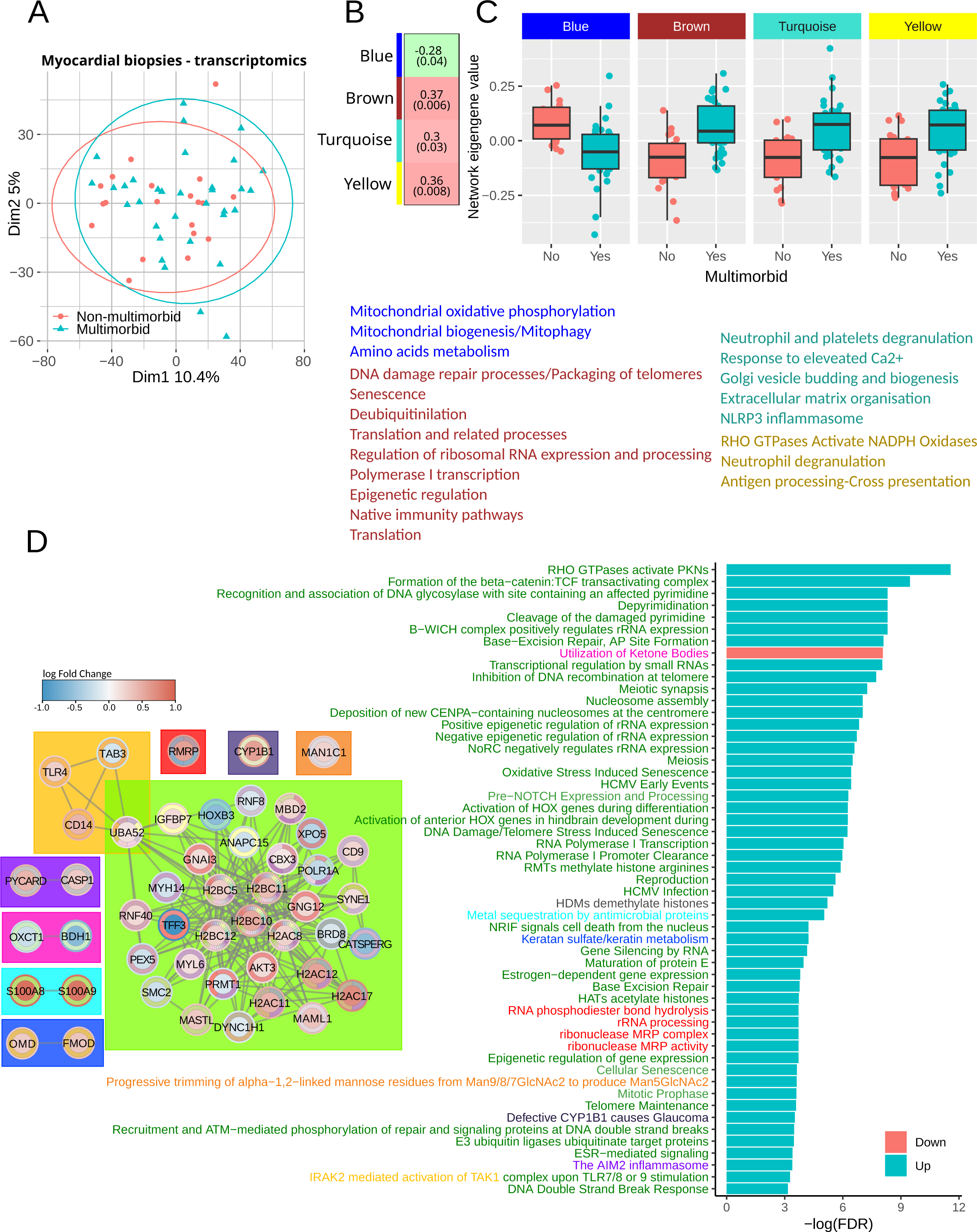
Transcripts analysis. **A** - Principal component (1 and 2) plots for the myocardial transcriptomics profiles. **B** – Transcripts networks identified in weighted gene correlation network analysis. Green indicates a negative correlation with the number of comorbidities, and red indicates a positive correlation. The numbers show a correlation estimate (p-value). **C** – Network eigengene values distribution between the groups. Genes constituting to each network were subjected to pathway enrichment analysis, and the summary is shown below the plots. The full list of pathways is in **Table S6**. **D** – Outcome of gene set enrichment analysis. The bar plot shows significantly regulated pathways (adjusted p-value<0.05) specifically in multimorbidity but not the comorbidities The full list in **Table S7**. The diagrams show transcripts sharing membership in at least one pathway. The node fill color shows transcripts’ log fold change with positive numbers indicating upregulation in multimorbidity. The colors in the node border indicate a number of pathways the transcript participates in.

Since none of the transcripts passed the multiple comparison adjustment, we first performed a weighted gene correlation analysis to identify significantly changing groups of transcripts. The analysis identified one network of transcripts that was significantly downregulated (blue in **Figure 3B, C** and **Table S5**) in the multimorbid myocardium and three significantly upregulated networks (brown, turquoise and yellow in **Figure 3B, C** and **Table S5**). The blue network included transcripts that enriched pathways involved in mitochondrial oxidative phosphorylation, mitochondrial biogenesis, mitophagy and amino acids metabolism. It included, among others, NUDS1, NUDV2, PDHB and OGHD. The brown network’s transcripts significantly enriched pathways involved in DNA damage processing and packaging, senescence, translation and regulation of ribosomal RNA expression. The transcripts included in this network were mainly ribosomal proteins, histone 2B and histone ubiquitin ligases. The turquoise and yellow networks included transcripts that significantly enriched innate immunity pathways. It included proteins required for neutrophil degranulation (LAMP1, PYCARD, FTL or RAB5C) but also proteins modulating inflammatory responses like serpins (A1, B1, E2, F1 and H1) and anti-apoptotic proteins like S100 (A4, A8, A9 and A11) or HMOX1. Transcript membership in each network is indicated in **Table S5,** and pathway enrichment results are in **Table S6**.

We also performed gene set enrichment analysis using the whole gene expression dataset (**Table S7**). The bar plot in **Figure 3D** shows pathways specific to multimorbidity that passed false discovery rate adjustment. Diagrams in the left panel show transcripts annotated to and linked by joint pathway membership. The significant transcripts involved in the enriched pathways mainly encode proteins regulating chromatin condensation and DNA repair: Ubiquitin-60S ribosomal protein L40 (UBA52), E3 ubiquitin-protein ligases RNF40, RNF8 and ANAPC15, Protein arginine N-methyltransferase 1 (PRMT1), Bromodomain-containing protein 8 (BRD8) and Serine/threonine-protein kinase greatwall (MASTL), Repressor methyl-CpG-binding domain protein 2 (MBD2), Chromobox protein homolog 3 (CBX3), Structural maintenance of chromosomes protein 2 (SMC2), Mastermind-like protein 1 (MAML1); transcripts involved in gene transcription and polymerase I-interacting proteins: DNA-directed RNA polymerase I subunit RPA1 (POLR1A), Homeodomain-containing DNA binding protein 3 (HOXB3); and transcripts encoding nuclear export and cytoskeletal proteins: Exportin-5 (XPO5), Myosin-14 (MYH14), Myosin light polypeptide 6 (MYL6), Nesprin-1 (SYNE1). The only downregulated pathway was the utilization of ketone bodies with two significantly downregulated transcripts: Succinyl-CoA:3-ketoacid coenzyme A transferase 1 (OXCT1) and D-β-hydroxybutyrate dehydrogenase (BDH1).

Inflammation-related pathways were upregulated in multimorbidity and all comorbidities but stroke. Mitochondrial electron transport chain, translation, protein import and biogenesis were also non-specific to multimorbidity and were downregulated in all comorbidities (**Table S7**).

To assess the influence of each comorbidity, we analyzed gene set enrichment without patients with that comorbidity. Pathways upregulation and downregulation patterns were similar across all comparisons. Removing patients with angina or previous MI had the strongest effect on the affected pathways (37% pathway overlap with the full dataset), as shown in **Figure S4**. However, it also considerably reduced the number of samples (13 multimorbid and 6 non-multimorbid). Similar to simple multimorbidity, complex multimorbidity affected DNA damage repair, nucleosome assembly, rRNA expression and translation (60% pathway overlap with the full dataset). The results of the analysis without patients receiving anti-diabetic medications were 91% identical to the analysis of the full dataset.

### Respiration in circulating monocytes

To assess the influence of multimorbidity on mitochondrial function, we measured respiration in blood monocytes, which are considered a good sensor for metabolic stressors. (Kramer et al., 2014)

The analysis of oxygen consumption rate (OCR) identified pre-operative basal (without any drugs) and maximal (in the presence of FCCP) respiratory control ratios (RCR) as significantly downregulated in the multimorbid samples (**Figure 4A-B**). The RCR is a measure of mitochondrial coupling (see the Methods section for definitions), which links respiration to ATP synthesis. However, the RCR component measurements were not different between the groups. Therefore, we analyzed correlations of the number of comorbidities with OCR measurements. Significant correlations were found with baseline OCR and basal RCR (**Figure 4C**).

**Figure 4.**
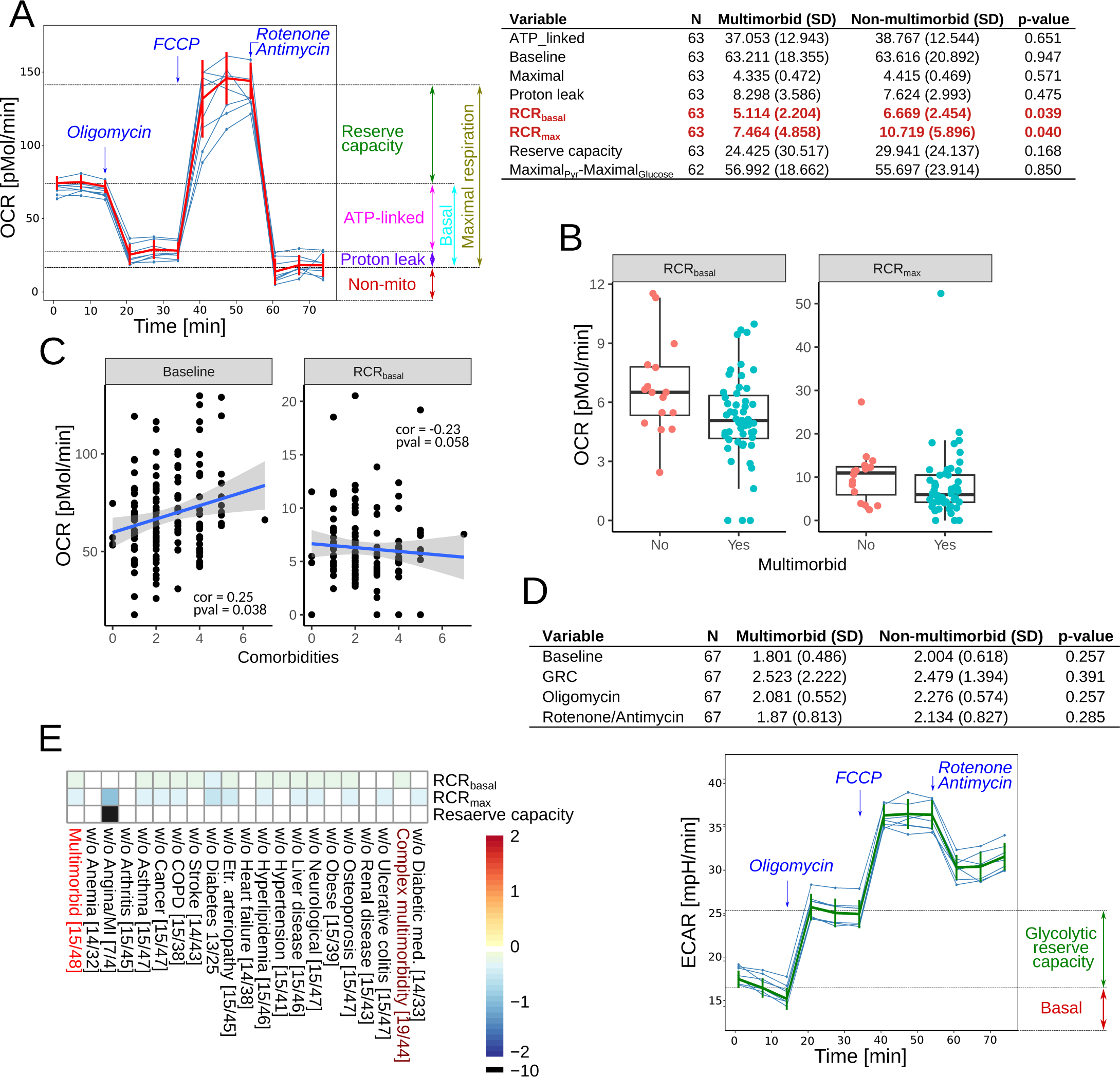
Mitochondrial respiration in monocytes. **A –** The assays were performed without or with mitochondrial toxins: oligomycin (ATPase synthase inhibitor), FCCP (drug uncoupling ATP synthesis and electron transport), antimycin A (complex III inhibitor) and rotenone (complex I inhibitor). The summary of the measured OCR parameters is shown in the left panel. The right panel shows the oxygen consumption rate in monocytes before and after surgery. Maximal_Pyr_-Maximal_Glucose_ is the difference between maximal respiration (in the presence of FCCP) in the presence of pyruvate or glucose. **B** – Box plots of the significantly different mitochondrial parameters between multimorbid and non-multimorbid samples. **C** – Plots of mitochondrial parameters significantly correlating with the number of comorbidities. **D** – ECAR in monocytes before and after surgery (upper panel) and summary of the measured ECAR parameters. **E** – Analysis of OCR parameters in complex multimorbidity and in reduced datasets to test the influence of each comorbidity and anti-diabetic medications. Color scale shows log fold change, where positive numbers indicate higher levels in multimorbidity. Numbers in square brackets indicate the number of non-multimorbid/multimorbid patients.

The analysis of extracellular acidification rate (ECAR), which is a measure proportional to glycolysis, did not find any significant differences (**Figure 4D**). To further test the role of glycolysis, we measured the difference in maximal OCR in the presence of glucose (glycolysis substrate) and pyruvate (glycolysis product). There was no difference in that measure, as well (**Figure 4A**, Maximal_Pyr_-Maximal_Glucose_).

To test the effect of the component comorbidities, we analyzed our dataset without patients with these comorbidities. Basal and maximal RCR decreased in most comparisons, except after removing patients with anemia, arthritis, heart failure or renal disease. Patients with complex multimorbidity had lower basal RCR, and patients not receiving anti-diabetic medications had lower maximal RCR. Removing patients with angina from the dataset resulted in decreased maximal RCR and reserve mitochondrial capacity. However, it also considerably reduced the dataset (multimorbid n=4, non-multimorbid n=7, **Figure 4E**).

### Energy metabolism in multimorbidity

Since we could not directly measure respiration in myocardial biopsies, we analyzed a set of metabolites involved in energy metabolism. Out of 144 metabolites, α-ketoglutarate, ATP, UTP, long-chain acyl-carnitines and formyl-valine significantly increased in multimorbidity, and NADH/NAD^+^ ratio decreased (**Figure 5A**). Further correlation analysis confirmed the role of α-ketoglutarate, ATP, UTP and formyl-valine, whose levels decreased with the number of comorbidities. Conversely, the NADH/NAD+ ratio increased with the number of comorbidities. In addition, the analysis identified cytosine, cytidine, and hydroxylated stearoylcarnitine (C18-OH) as positively correlated and asymmetric dimethylarginine (ADMA) as negatively correlated with the number of comorbidities (**Figure 5B**).

**Figure 5.**
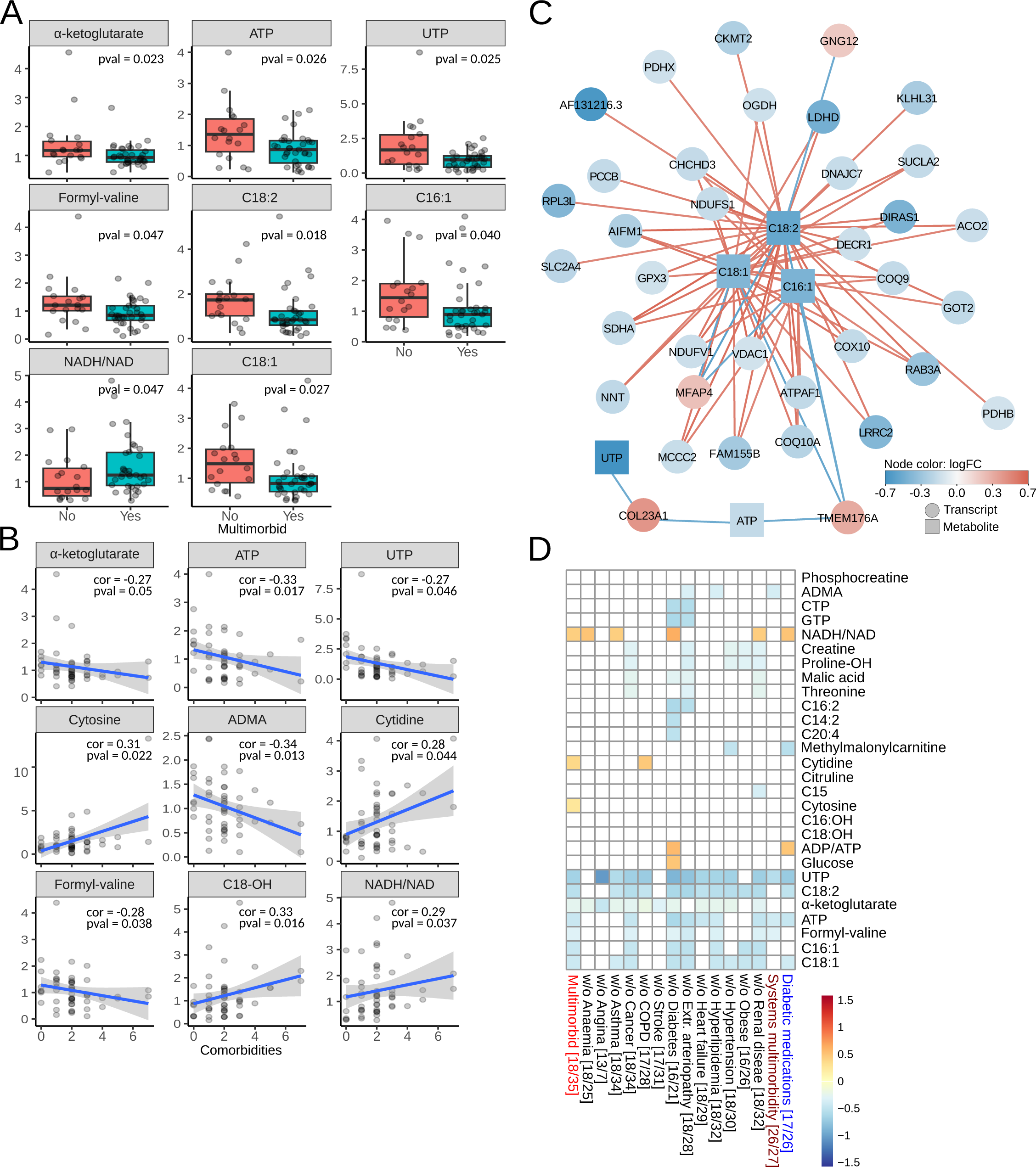
Metabolites involved in energy metabolism. **A** – Targeted metabolites significantly different in multimorbid myocardium. **B** - Targeted metabolites significantly correlating with the number of comorbidities. **C** – Transcripts and metabolites were combined using sparse Partial Least Squares models with 0.6 cutoff. Node color indicates expression log fold change in the multimorbid group. Red edges show positive and blue edges show a negative correlation between nodes. **D** – Analysis of targeted metabolites in complex multimorbidity and in reduced datasets to test the influence of each comorbidity and anti-diabetic medications. Color scale shows log fold change, where positive numbers indicate higher levels in multimorbidity. Numbers in square brackets indicate the number of non-multimorbid/multimorbid patients.

The metabolites, which were significant in group-wise comparisons and correlation analysis, were combined with paired transcriptomics in weighted gene correlation analysis using correlation cutoffs above and below 0.6 and −0.6, respectively. The three long-chain acyl-carnitines (C18:1, C18:2 and C16:1) positively correlated mainly with transcripts involved in the citric acid cycle and respiratory electron transport chain: NUDS1 NUDV2, PDHB, OGHD, COQ10A, ACO2, SUCLA2, NNT, PDHX, SDHA, VDAC1 but also with solute carrier SLC2A4 and heart-specific ribosomal protein RPL3L. A negative correlation was found between Guanine nucleotide-binding protein subunit gamma-12 (GNG12), Microfibril-associated glycoprotein 4 (MFAP4) and Transmembrane protein 176A (TMEM176A). ATP and UTP correlated negatively with TMEM176A and Collagen α-1 (XXIII) chain (**Figure 5C**).

The influence of single comorbidities was analyzed in datasets without these comorbidities. **Figure 5D** shows that ATP, UTP, α-ketoglutarate, and long-chain acyl-carnitines were decreased in most of the analyses. That was also true for a dataset without patients on anti-diabetic medications. Patients with complex multimorbidity had decreased levels of ATP, UTP, long-chain acyl-carnitines and ADMA.

### Senescence-associated secretory phenotype

Decreased levels of energy substrates and nucleoside triphosphates can indicate higher levels of senescence. This is supported by changes in senescence-associated processes and pathways, including downregulation of mitochondrial oxidative respiration and mitochondrial biogenesis and upregulation of inflammation or DNA damage response. Transcripts like IGFBP7 (**Figure 3D**), TIMP1, collagen, laminin, CCL13 and C-C/C-X-C chemokines receptors (**Table S5**), showing differential regulation in the multimorbid myocardial samples, encode proteins whose expression changes in senescence and are part of the senescence-associated secretory phenotype (SASP). (Coppé et al., 2010; Dookun et al., 2020) Therefore, we tested whether cytokines previously described as SASP proteins were upregulated in the pre-operative plasma samples. To do this, we analyzed a panel of 71 cytokines and chemokines in samples from 48 participants. The panel includes cytokines that are regulated by NF-κB or IL-1/NLRP3, which are major modulators and initiators of SASP expression. (Acosta et al., 2013; Salminen et al., 2012)

Groupwise comparison indicated that fractalkine and IL-22 were significantly upregulated in the multimorbid samples (**Figure 6A**). Further correlation analysis with the number of comorbidities added GM-CSF, IL-1β, IL-1RA and IL-3 to the list (**Figure 6B**). Downregulation of both fractalkine and IL-22 was not affected by any comorbidity or anti-diabetic medications, as indicated by the analysis in reduced datasets. Both proteins were also downregulated in complex multimorbidity (**Figure 6D**).

**Figure 6.**
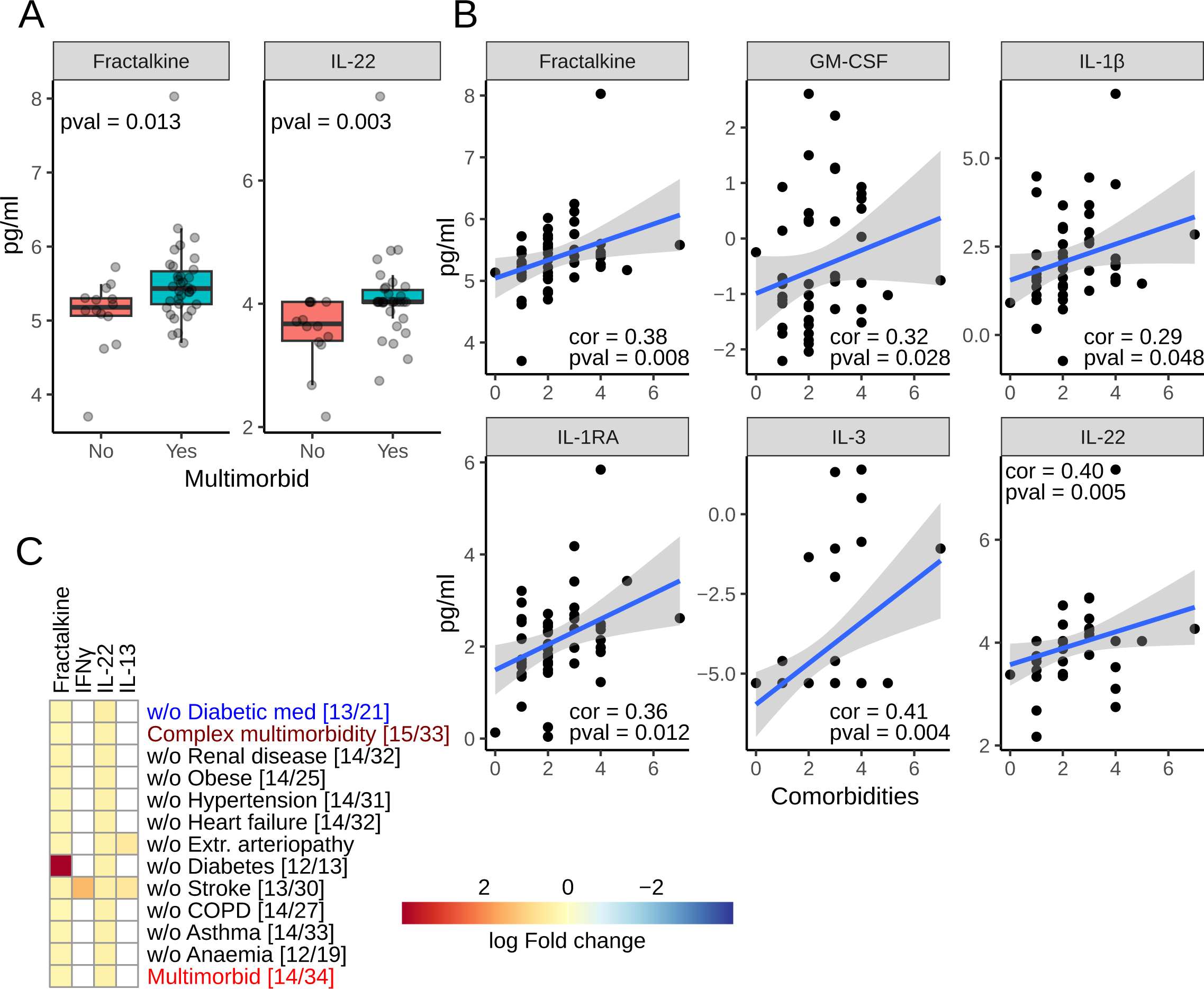
Senescence-associated secretory phenotype. **A –** Box plots of analytes significantly different in multimorbidity. **B** – Plots of analytes correlating significantly with the number of comorbidities. **C** – Analysis of the analyte panel in complex multimorbidity and in reduced datasets to test the influence of each comorbidity and anti-diabetic medications. The color indicates log fold change in multimorbidity. Numbers in square brackets indicate the number of non-multimorbid/multimorbid patients.

### DNA damage and nucleolar assembly

Transcriptomics data suggested that the primary driver of senescence, DNA damage, is upregulated in multimorbidity. In addition, several transcripts encoding ribosomal proteins and others involved in polymerase I transcription and ribosomal RNA processing were affected (**Figure 3C-D**). This can indicate changes in the expression of ribosomal RNA and dysregulation of nucleolar assembly, which can result in free ribosomal proteins, increased senescence levels and, consequently, changes in mitochondrial respiration. (W. Wang et al., 2015)

We first tested ribosomal DNA (rDNA) copy numbers, as it was previously linked with mitochondrial abundance. (Gibbons et al., 2014) We analyzed rDNA in myocardial biopsies with qRT-PCR and specific primers, and as shown in **Figure 7A**, we did not detect any difference in rDNA copy numbers between the groups. DNA damage was detected with antibodies against the phosphorylated form of histone 2AX (γH2AX) in myocardial cryo-slices. Multimorbid samples had significantly more nuclei positive for γH2AX (**Figure 7B, C**). Next, nucleolar stress was assessed with antibodies against nucleolin and fibrillarin. Although fibrillarin staining patterns were not different in multimorbid samples, nucleolin labeling suggested higher levels of nucleolar stress as indicated by a larger fraction of nuclear area positive for nucleolin (**Figure 7D – F**).

**Figure 7.**
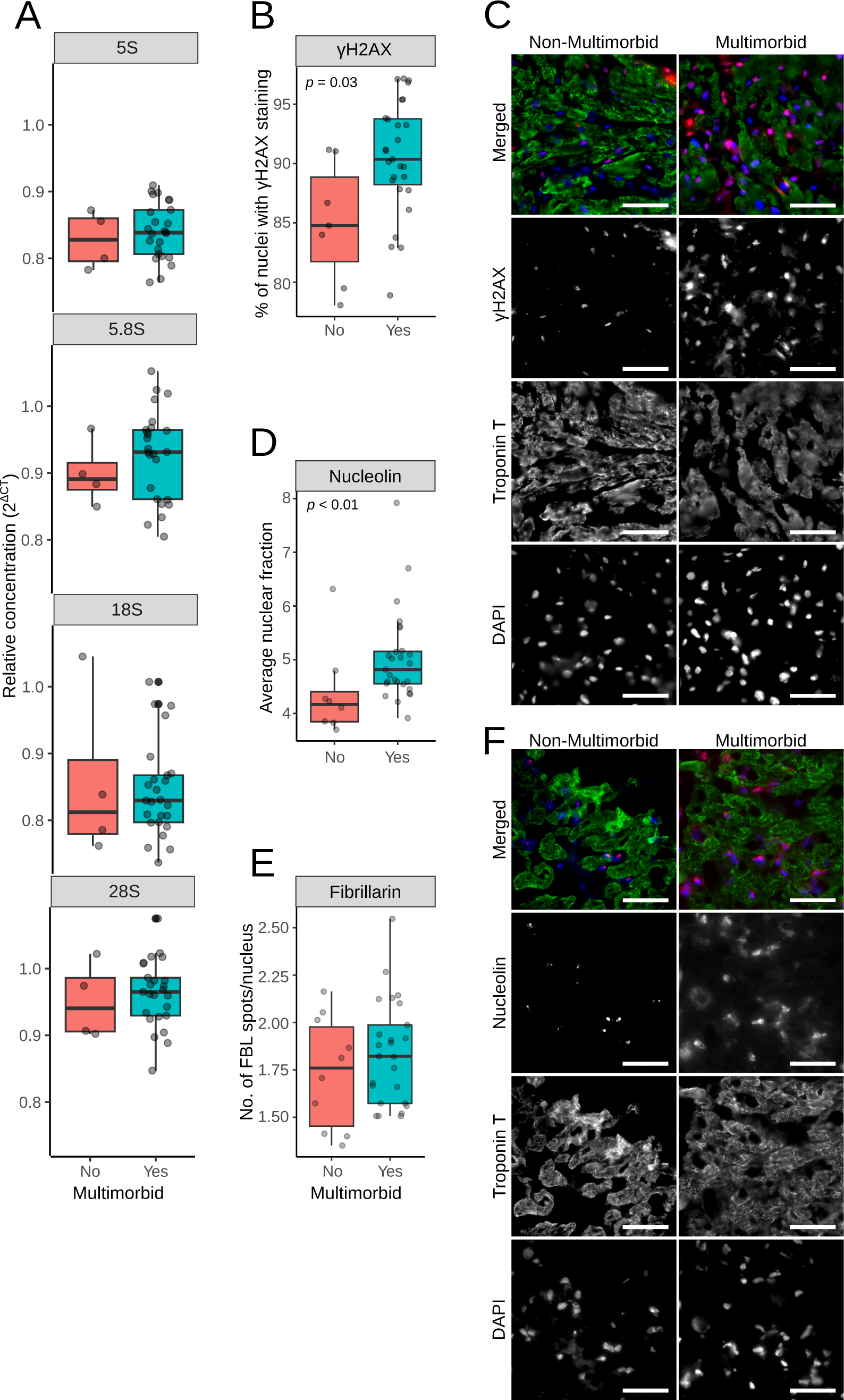
DNA damage and nucleolar stress. **A –** Ribosomal DNA copy numbers were analyzed by qRT-PCR with specific primers to genes encoding ribosomal DNA. The CT values were normalized against β-2-Microglobulin. **B** – γH2AX was detected in cryosections of myocardial biopsies, and the numbers indicating the percentage of γH2AX-positive cells were plotted. **C** – Representative images of cryosections labelled with γH2AX. D – Nucleolin was detected in cryosection, and the plots show the percentage of nuclear area occupied by the nucleolin staining. **E** – Representative images after the nucleolin labelling. Scale bars in C and E are 50 µm. **F** – Fibrillarin was detected in cryosection, and plots show the number of nuclear fibrillarin spots.

## Discussion

Using unbiased untargeted approaches, we identified processes specific to multimorbidity in people with cardiovascular disease that are characteristic of biological ageing and senescence. In the myocardium, these include changes at the transcriptomics level in chromatin organization pathways, DNA damage repair, nucleolar function and ketone bodies’ utilization in mitochondria. At the metabolite level, we observed decreased levels of products and substrates of mitochondrial oxidative phosphorylation. Mitochondrial function was also affected at the systemic level, as indicated by basal respiration that increased with the number of comorbidities. Patients with multimorbidity also had increased levels of caffeine, its breakdown products and intermediates of bile acid biosynthesis. Further validation experiments confirmed that senescence is associated with multimorbidity, as levels of several proteins of senescence-associated secretory phenotype increased with the number of comorbidities. We also confirmed that increased DNA damage and nucleolar instability play a role in multimorbidity.

### Clinical significance

Our transcriptomics analysis identified three major processes that are affected by multimorbidity. These are immune system activation, epigenetic regulation of gene expression and DNA repair, and mitochondrial energy production.

The specifically enriched epigenetic pathways include histones and enzymes mediating post-translational histone modifications. The expression of histone 2B (H2B) and its E3 ubiquitin ligase (RNF40) and several other ubiquitin ligases were increased in the multimorbid group. H2B ubiquitination on K120 stimulates rapid changes in chromatin remodeling and transcriptional activity mediated by p53, a transcription factor involved in senescence. (Minsky et al., 2008) Although we did not observe upregulation of CDKN1A (human p21 orthologue), several genes regulated by TP53 (human p53 orthologue) were affected. These include IGFBP7, TAB3 (TGF-β-activated kinase 1), AKT3 (RAC-gamma serine/threonine-protein kinase), ANAPC15, GNG12 (G protein subunit gamma 12), (Fischer, 2017) all of which are implicated in senescence. Dysregulation of the epigenetic regulation is also known to destabilize nucleolar function and ribosomal assembly, resulting in free ribosomal proteins that trigger senescence and alter DNA repair. (W. Wang et al., 2015) For example, free cytoplasmic ribosomal proteins L23, L29, S3 and S15 (upregulated in our dataset) bind MDM2, a TP53 ubiquitin ligase, enabling senescence-specific gene expression. (W. Wang et al., 2015) Free nuclear RPS3 binds to oxidative lesions in DNA and inhibits DNA repair, (Hegde et al., 2007) and RPL3 upregulates CDKN1A inducing mitochondrial-driven apoptosis. (Russo et al., 2013)

Transcriptional changes demonstrated enrichment of senescence-linked pathways, which was confirmed by analysis of cytokines and chemokines in plasma. Several members of the senescence-associated secretory phenotype (IL-1β, IL-1RA and GM-CSF (Coppé et al., 2010)) correlated positively with the number of comorbidities. Fractalkine, whose levels increased in the multimorbid group and correlated with the number of comorbidities, was previously reported to decrease in a mouse model after senolytic treatments. (Dookun et al., 2020) Levels of IL-22 showed a similar pattern to fractalkine. IL-22 is released by T cells as part of the tissue repair response, (Dudakov et al., 2015) and its overexpression in hepatic cells induced senescence in a p53 and p21-dependent manner. (Kong et al., 2012)

Caffeine and its breakdown products accumulated in the heart tissue and plasma before and after surgery. It is normally broken down in the liver’s endoplasmic reticulum by cytochrome p450 enzymes, followed by further processing in lysosomes before secretion in urine. The effect could be a consequence of a changed NADH/NAD+ ratio, which influences the activity of cytochrome p450 enzymes and, consequently, oxidation of fatty acids and steroids, whose expression changes were also observed in plasma. Cytochrome p450 enzymes are often affected in polypharmacy and are closely associated with aging. (Doan et al., 2013) Several of them showed decreased expression in the myocardium in the multimorbid group.

The processes observed in people with multimorbidity are consistent with the ‘hallmarks of biological ageing’, a cluster of cellular changes implicated in the pathogenesis of age-related diseases. (López-Otín et al., 2023) Importantly, these processes are targets for novel and established therapeutics. For example, the administration of nicotinamide riboside can modulate the ageing process, (D. D. Wang et al., 2022) and the administration of sodium-glucose cotransporter 2 inhibitors alters nutrient sensing, mitochondrial dysfunction and autophagy in heart failure. (Packer, 2022) In addition, more established drugs like metformin or rapamycin that modulate mTOR could be effective in reversing DNA damage and unfavourable epigenetic changes.

### Strengths and limitations

The analysis has several strengths. First, the hypothesis was tested a priori using unbiased techniques. Second, the results of the untargeted analyses were consistent between omics platforms and were reproduced across different targeted assays. Our findings also agree with contemporary reviews of the pathogenesis multimorbidity. Third, to our knowledge, this is the first evaluation of mechanisms underlying multimorbidity in human myocardium. Almost all of the existing evidence is derived from animal studies or analyses of human blood.

Important limitations are: First, the small sample size and the breadth of the data increase the likelihood of statistical error in our analyses. In addition, myocardial biopsies may not have been uniform in cell content and included fragments of blood vessels or fat tissue, which further adds to the heterogeneity between samples. Due to the small sizes of the biopsies, it was impossible to estimate the variation in the cell composition. In mitigation, fluorescent image analysis indicates that most cells were cardiomyocytes. However, we cannot exclude the possibility that cells other than cardiomyocytes reduced differences in the multimorbid group, as different cell types differ in gene expression in response to multimorbidity. (Tian et al., 2023) Further studies, taking advantage of single-cell technologies, which can separate cells *in silico,* will help better define differences in the multimorbid group depending on the cell type. Second, we used a widely used consensus definition of multimorbidity. (I. S. S. Ho et al., 2022; Quality standard [QS153], 2017) This definition is limited because it is likely that clusters of chronic conditions not reflected in this definition lead to specific multimorbidity phenotypes. However, comparison to the complex multimorbidity, where two or more physiological systems are affected, generally supported our initial findings. The results also remained consistent when testing the influence of diabetic drugs (significantly enriched in the multimorbid group) or the underlying comorbidities. Most influential was angina or previous MI, likely because most patients had this comorbidity and removing them considerably reduced our dataset. It reflects the nature of our cohort, which was enriched with atherosclerosis risk factors and conditions typical of a low-risk cardiac surgery population. Therefore, our findings are limited to multimorbid patients with cardiovascular disease and may not apply to a wider population of people with multimorbidity. Moreover, the recruited cohort was at low risk even for cardiac surgery; only 8% of participants developed acute kidney injury, whereas >25% is commonly observed in unselected cardiac surgery populations. (Karkouti et al., 2009) This resulted in only small differences in adverse outcomes post-surgery between the multimorbid and non-multimorbid groups and was likely a factor that prevented us from detecting significant changes at transcript and metabolite levels in the untargeted analyses. A high-risk surgery cohort, or multimorbidity in older, sicker patients presenting with ACS, may have resulted in a stronger effect size. The fact that almost all changes observed in the multimorbid group were positively associated with the number of comorbid conditions supports this conjecture. Furthermore, the cardiac surgery population is heterogeneous and larger epidemiological studies that cluster patients into specific phenotypes will better characterize mechanisms of multimorbidity. Third, we excluded patients with pre-existing paroxysmal, persistent or chronic atrial fibrillation or pre-existing inflammatory states. These are well-defined conditions with characteristic phenotypes and a strong effect size that could introduce important heterogeneity in a small sample. In mitigation, these comorbidities are extremely rare, and the recruited cohort is typical of the normal adult cardiac surgery population.

## Conclusion

A prospective multi-omics analysis of human blood and myocardium obtained from a cohort undergoing cardiac surgery has identified multiple hallmarks of biological aging associated with multimorbidity, with epigenetic changes and cellular senescence being specific. Many of these processes, including mitochondrial dysfunction, chronic inflammation, and cell senescence, are modifiable by existing treatments.

## Author Contributions

MJW and GJM designed the study; MJW, GJM, KT and MR wrote the manuscript and prepared tables and figures; MR, HA and LJ-D managed the conduct of the study; FYL, MR, HA and LJ-D managed the data during the study; KT, MJW, SY, BE-H and SS collected samples and undertook the laboratory analyses; MG, AM and JLG performed targeted metabolite analysis. MJW and FYL carried out statistical analyses; MJW and ASA performed bioinformatics analyses. GDR shared experimental protocols and contributed to paper writing. All authors reviewed the report for important intellectual content and approved the final version.

## Data Availability

Sequencing data are available via NCBI Gene Expression Omnibus (GSE159612). Metabolomics data are available through EMBL-EBI (MTBLS7259).

---

**Figure S1.**
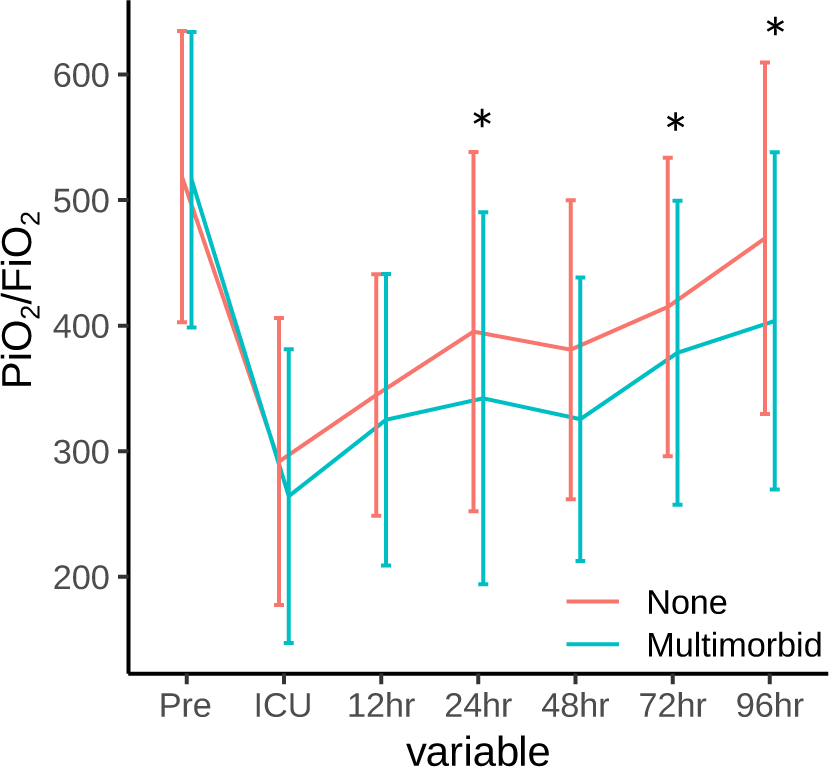
- PiO2/FiO2 ratio before and after surgery. Asterisks indicate a significant difference (p-value < 0.05)

**Figure S2.**
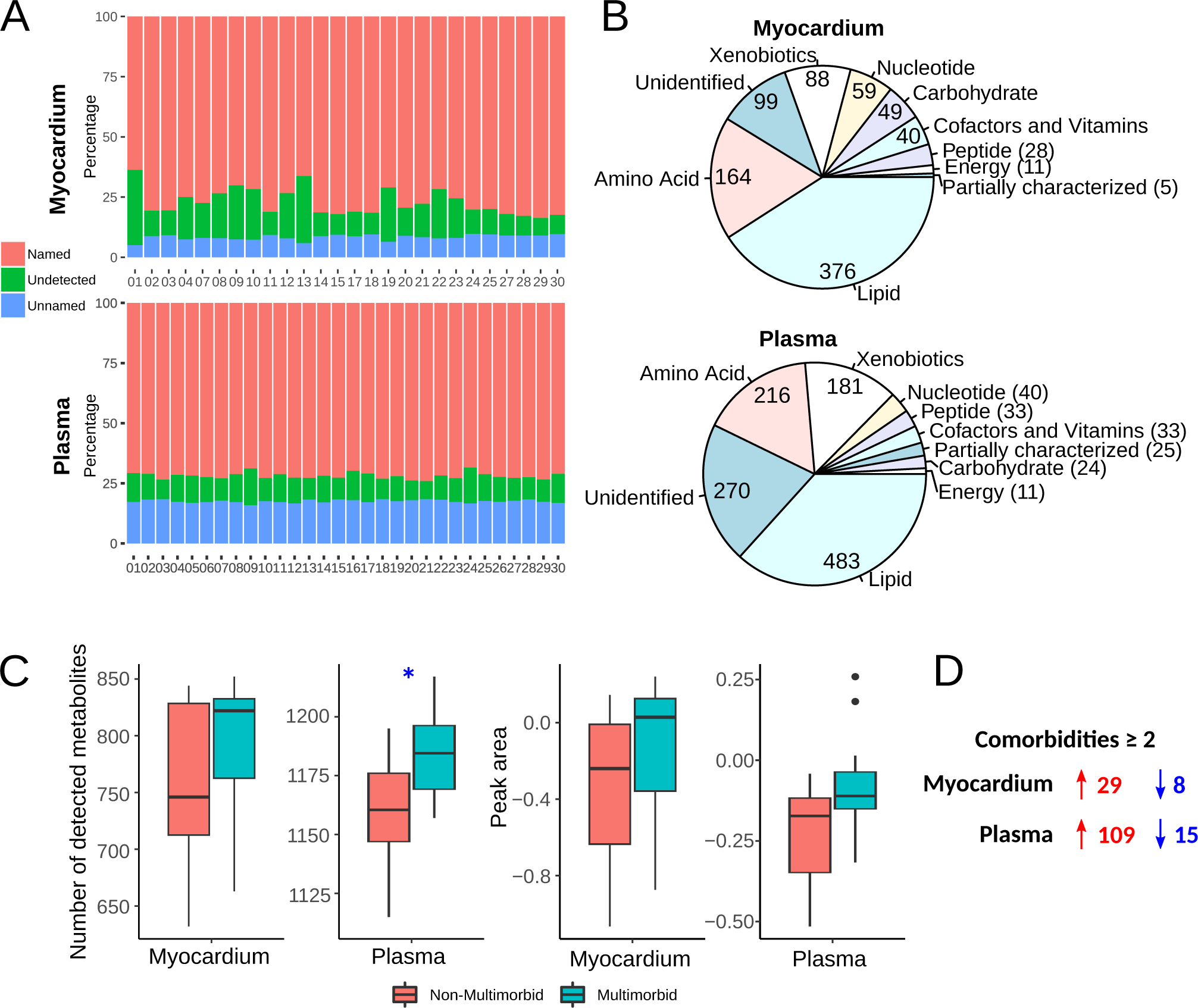
– Metabolomics summary. **A** – Distribution of named, unknown and undetected metabolites in all sample types. **B** – Metabolites biotype distribution in all sample types. **C** – Comparison of detected metabolite numbers and average peak ratio in all sample types. **D** – summary of the differential expression analysis.

**Figure S3.**
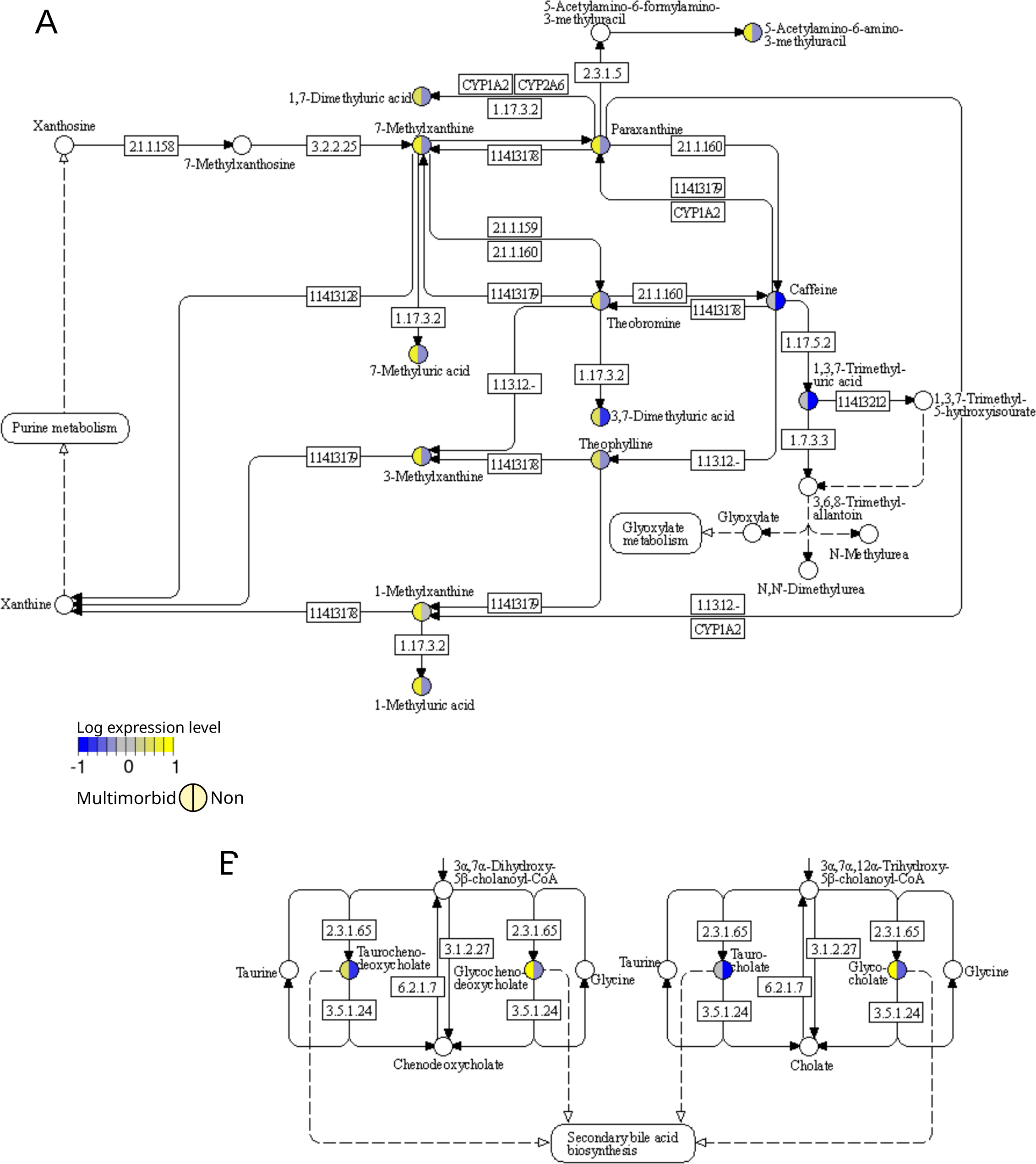
– Pathway overlay. Expression of significantly different metabolites with membership in KEGG’s Caffeine metabolism **(A)** and Primary bile acid biosynthesis **(B)** pathways.

**Figure S4.**
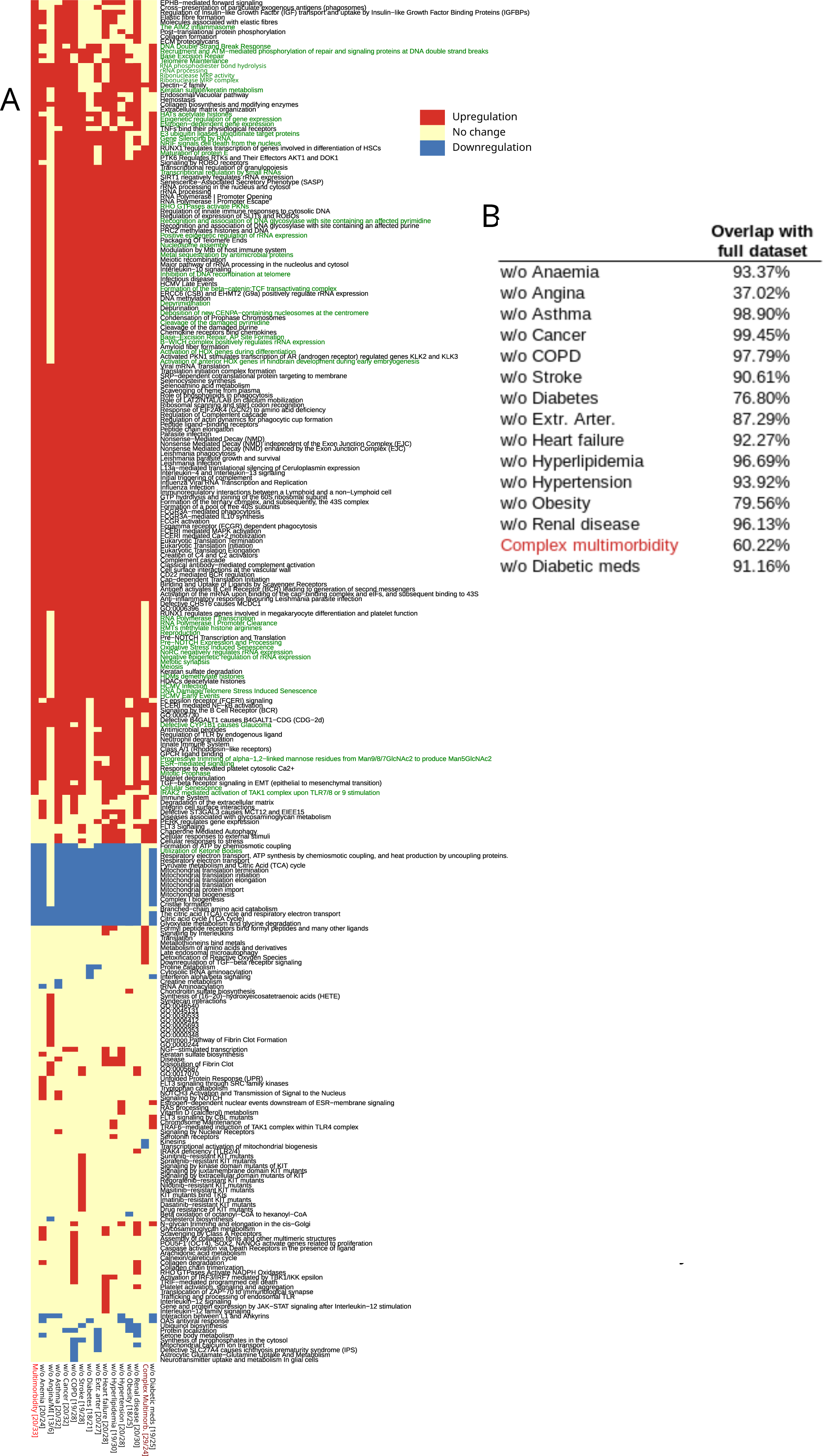
**– A** - Gene set enrichment analysis in complex multimorbidity and in reduced datasets to test the influence of each comorbidity and anti-diabetic medications. Numbers in square brackets indicate the number of non-multimorbid/ multimorbid patients. **B** - Pathway overlap with the full dataset for each analysis.

